# Autosomal recessive *SLC30A9* Mutations in a Proband with a Cerebro-Renal Syndrome and No Parental Consanguinity

**DOI:** 10.1101/2021.07.21.21260807

**Authors:** Robert Kleyner, Arif Mohammad, Elaine Marchi, Naomi Horowitz, Andrea Haworth, Brian King, Maureen Gavin, Karen Amble, Milen Velinov, Gholson J. Lyon

## Abstract

An *SLC30A9-*associated cerebro-renal syndrome was first reported in consanguineous Bedouin kindred by Perez et al. in 2017. While the function of the gene has not yet been fully elucidated, it may be implicated in Wnt signaling, nuclear regulation, as well as cell and mitochondrial zinc regulation. In this research report, we present a female proband with two distinct, inherited autosomal recessive loss-of-function *SLC30A9* variants from unrelated parents. To our knowledge, this is the first reported case of a possible *SLC30A9-*associated cerebro-renal syndrome in a non-consanguineous family. Furthermore, a limited statistical analysis was conducted to identify possible allele frequency differences between populations. Our findings provide further support for an *SLC30A9*-associated cerebro-renal syndrome and may help further clarify the gene’s function.

## Introduction

An autosomal recessive cerebro-renal syndrome associated with mutations in gene *SLC30A9* was first reported by Perez et al. in a consanguineous Bedouin family in 2017. While the clinical manifestations of the syndrome are variable, all six individuals investigated had decreased renal function, developmental delays, truncal hypotonia, ataxia, spasticity, camptocormia, and hypertonia of limb muscles.

*SLC30A9* is a member of the SLC30 family of zinc transporters (ZnT) responsible for maintaining zinc homeostasis by transporting zinc from the cytosol to organelles and the extracellular space to avoid toxicity (Palmiter and Huang 2004). SLC30 ZnTs have been previously implicated in transient neonatal zinc deficiency, diabetes mellitus, hepatic cirrhosis, polycythemia, hypermagnesemia, dystonia, and parkinsonism (Kambe, Hashimoto, and Fujimoto 2014). Some ZnT proteins have also been implicated in pancreatic, breast, and prostate cancers (Bafaro et al. 2017).

In this case study, we describe a female proband with two distinct, inherited loss of function mutations in *SLC30A9* presenting with clinical findings similar to those described by Perez et al. To our knowledge, this is the first known case of a possible *SLC30A9-*associated cerebro-renal syndrome in a non-consanguineous family. The variants were identified through next-generation whole exome sequencing (WES) and were validated using the Sanger sequencing method. This case study may provide further support for an *SLC30A9-*associated cerebro-renal syndrome and may offer more insight in the role of *SLC30A9* in metabolism and human disease.

### Clinical Presentation

The proband first presented to our clinic and agency for services evaluation from the Office of People with Developmental Disabilities in New York State at around one year of age, after referral for microcephaly and developmental delay. A comparison of the clinical features found in the proband and Bedouin kindred reported by Perez et al. is included in **Table 1**. An extensive clinical summary is included in **Supplementary Information**. The family signed informed consent to participate in research and to have the results published.

**Table 1:** Summary of clinical findings found in the proband, as well as those in the Bedouin kindred described by Perez et al.

| Clinical Feature | Proband | Bedouin Kindred |
| --- | --- | --- |
| **Cardiac** |  |  |
| Heart murmur (HP:0030148) | - | N/A |
| **Craniofacial** |  |  |
| Abnormality of cranial sutures (HP:0011329) | - | N/A |
| Craniosynostosis (HP:0001363) | - | - |
| Facial hypotonia (HP:0000297) | + | - |
| Long eyelashes (HP:0000527) | + | - |
| Upslanted palpebral fissure (HP:0000582) | + | - |
| **Developmental** |  |  |
| Failure to thrive in infancy (HP:0001531) | + | - |
| Feeding difficulties (HP:0011968) | + | N/A |
| Growth delay (HP:0001510) | + | N/A |
| Severe global developmental delay (HP:0011344) | + | + |
| **Musculoskeletal** |  |  |
| Appendicular hypotonia (HP:0012389) | - | - |
| Muscular hypotonia of the trunk (HP:0008936) | + | + |
| **Neurological** |  |  |
| Abnormal brainstem morphology (HP:0002363) | - | - |
| Abnormal cerebellum morphology (HP:0001317) | - | - |
| Abnormal cerebral vascular morphology (HP:0100659) | - | - |
| Abnormal cerebral ventricle morphology (HP:0002118) | - | - |
| Abnormal cerebral white matter morphology (HP:0002500) | + | - |
| Abnormal CNS myelination (HP:0011400) | - | - |
| Abnormality of the pituitary gland (HP:0012503) | - | - |
| Agenesis of corpus callosum (HP:0001274) | + | - |
| Arachnoid cyst (HP:0100702) | + | - |
| Gray matter heterotopia (HP:0002282) | - | - |
| Hydrocephalus (HP:0000238) | - | - |
| Microcephaly (HP:0000252) | + | - |
| Pachygyria (HP:0001302) | + | - |
| Progressive sensorineural hearing impairment (HP:0000408) | + | N/A |
| **Obstetrical/Neonatal** |  |  |
| Abnormality of the abdominal organs (HP:0002012) | - | N/A |
| Intrauterine Growth Restriction (HP:0001511) | + | - |
| Neonatal respiratory distress (HP:0002643) | + | - |
| Two-Vessel Umbilical Cord (HP:0001195) | + | - |
| **Renal** |  |  |
| Abnormal renal morphology (HP:0012210) | - | + |
| Renal hypoplasia (HP:0000089) | + | - |
| Stage 3 chronic kidney disease (HP:0012625) | + | + |
| **Serum** |  |  |
| Abnormal circulating acetylcarnitine concentration (HP:0012071) | +/- | - |
| Increased circulating creatine kinase MB isoform (HP:0032232) | + | - |

### WES Analysis

Samples from the proband (around age range 5-10 years old) and two relatives were sent to Novogene (Sacramento, CA) for clinical WES. Variant calling and interpretation was performed using the Cogenica platform (Congenica Limited, Cambridge, UK). Coverage and mapping statistics for whole exome sequencing performed on the family are shown in **Supplementary Table 1**. Direct Sanger sequencing was performed to validate suspected mutations. The results of the sequencing supported the presence of both variants in the proband. Sanger sequencing chromatograms are presented in **Figure 1**. Methods are discussed at length in **Supplemental Information**.

**Figure 1:**
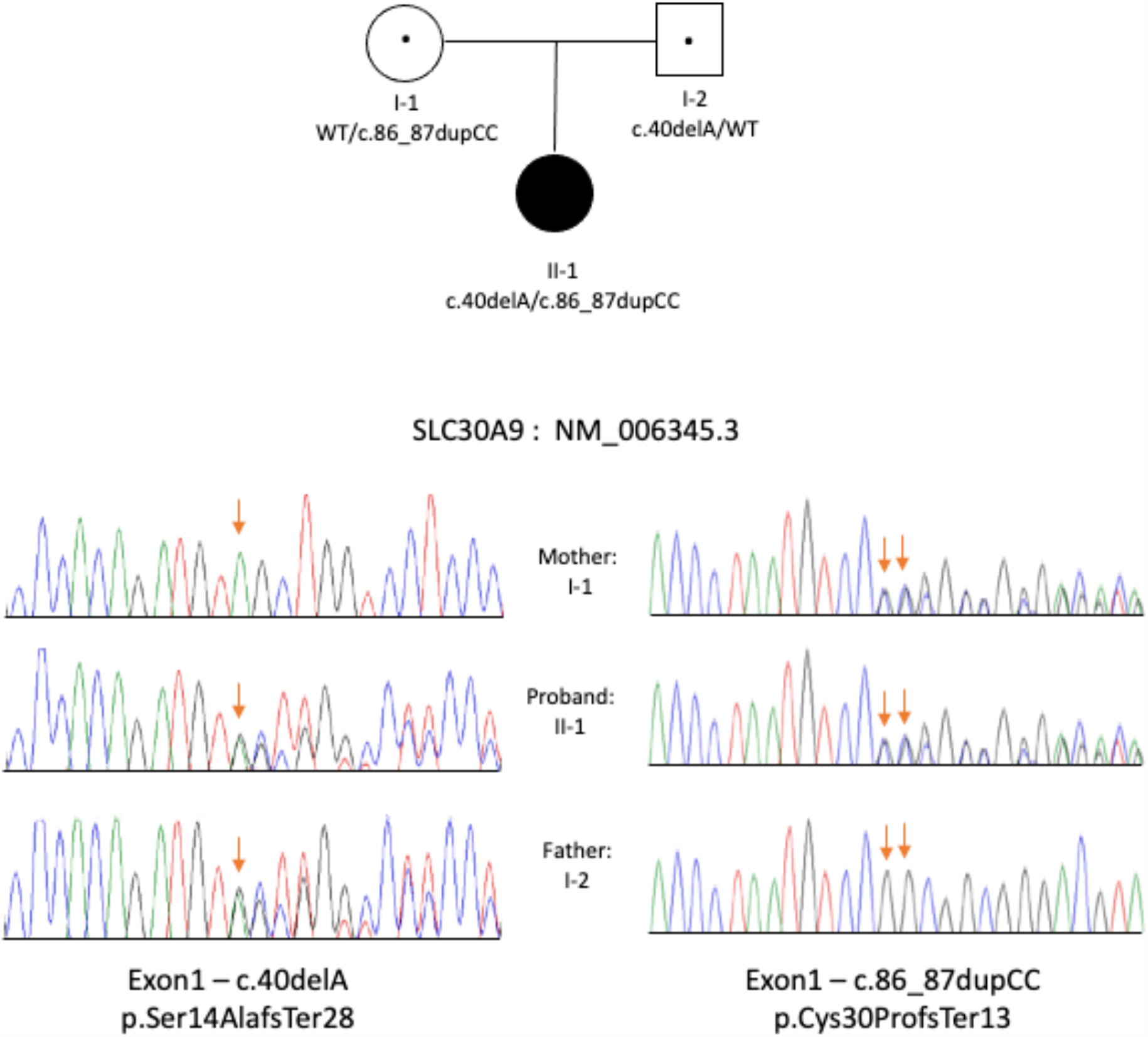
The proband (II-1) carried compound heterozygous frameshift mutations as confirmed by Sanger sequencing: c.40delA inherited from the father (I-2) and c.86_87dupCC from the mother (I-1).

### Population Analysis of *SLC30A9* Variants

Given that this proband was of African American descent, it was hypothesized that variants in *SLC30A9* may be more common in individuals with African ancestry. Pathogenic mutations in the gene have also been reported in a Bedouin kindred, who may possess African haplotypes (Abu-Amero et al. 2008). gnomAD was used to identify possible differences in heterozygous missense and loss of function (LoF) variant allele frequencies (AFs). AFs were computed for African/African American, Latino/Admixed American, Ashkenazi Jewish, East Asian, Finnish-European, Non-Finnish European, South Asian, and “Other” populations. “Other” populations are represented by individuals who did not cluster with the other populations after a principal component analysis. Given that gnomAD contains 76,156 genomes of individuals with no known medical or family history of severe pediatric disease, it is assumed that the heterozygous variants do not contribute to a cerebro-renal syndrome if present alone (Karczewski et al. 2020). Summary statistics are included in **Supplementary Table 3** and a PDF of a Python *Jupyter* notebook detailing this analysis is included as **Supplementary File 1**.

The African/African American, as well as the Latino/Admixed American, East Asian, and South Asian populations, had significantly greater missense *SLC30A9* variant AFs when compared to the Ashkenazi Jewish, Finnish-European, and “Other” populations. The calculated missense variant AFs in the non-Finnish European population were significantly greater when compared to the other seven populations analyzed. A heat map depicting these results is shown in **Supplementary Figure 1A**. Calculated AFs for heterozygous LoF mutations in *SLC30A9* were significantly greater in the African/African, East Asian, and South Asian populations than the “Other” and Ashkenazi Jewish populations. Consistent with the findings for missense AFs, the non-Finnish European population had significantly greater AFs than all other populations. These results are displayed as a heatmap in **Supplementary Figure 1B**.

## Discussion

In this report, we present a female proband who is compound heterozygous for two novel loss of function variants in *SLC30A9*. There are several similarities between her phenotype and the clinical features reported in a Bedouin family by Perez et al. We believe that our findings provide additional support for the existence of a *SLC30A9*-associated cerebro-renal syndrome, which in turn emphasizes the gene’s importance in zinc homeostasis.

Individuals with African Ancestry do appear to be more likely to carry missense and LoF mutations in *SLC30A9* variants when compared to individuals with Ashkenazi Jewish and “Other” gnomAD populations, although individuals with European ancestry appear to be most at risk.

Both experimental and observational studies have demonstrated the role of zinc metabolism in neurological diseases such as neurodegenerative disorders, autism spectrum disorder, movement disorders, amyotrophic lateral sclerosis, mood disorders, traumatic brain injury, strokes, and seizures (Prakash, Bharti, and Majeed 2015). Zinc released from neuron synapses plays a role in regulating GABA, glycine, N-methyl-D-aspartate (NMDA), and α- amino-3-hydroxy-5-methyl-4-isoxazole-propionate (AMPA) receptors (Smart, Hosie, and Miller 2004; Szewczyk 2013). Furthermore, zinc has been demonstrated to have a dose-dependent neuroprotective effect, with toxicity noticed at higher dosages (Choi et al. 2020). The SLC30 family of proteins has specifically been implicated in movement disorders and Alzheimer’s disease (Quadri et al. 2012; Xu et al. 2019).

Although the role of zinc transporters and homeostasis in renal pathophysiology is less characterized, several observational studies support the clinical relevance of zinc in renal disease. Individuals with CKD were found to have decreased concentrations of plasma and urinary zinc, as well as an increased fractional excretion of zinc when compared to healthy control subjects (Damianaki et al. 2020). Low serum zinc levels may also be associated with the progression of diabetic nephropathy, given that it was found to be inversely correlated with microalbuminuria and serum creatinine, and directly correlated with eGFR in diabetic individuals (Al-Timimi, Sulieman, and Hussen 2014). Analysis of data from the Korean Genome and Epidemiology Study suggests a possible causative relationship; individuals whose dietary zinc consumption was calculated to be in the first quartile had a 36% greater risk of developing CKD than those whose zinc intake was in the fourth quartile (Joo et al. 2021).

Experimental studies have mixed results in support for these findings. Zinc supplementation has been found to have a protective effect against gentamicin-induced nephropathy in rats (Teslariu et al. 2016). However, in children with CKD, zinc supplementation was found to have a significant positive change in body mass and resulted in “normalization,” but no significant change in serum albumin, zinc and CRP levels (Escobedo-Monge et al. 2019).

There are several limitations to our conclusions. Although we have made every effort to provide tangible clinical benefit to the proband and her family, clinic visits were sporadic, and the patient was eventually lost to follow-up. Consequently, it is likely that additional clinical features were not presented in this case report, as we were unable to obtain all medical records regarding the proband’s care.

Challenges with obtaining paternal DNA samples provide further uncertainty regarding the significance of a 9p24.1-p24.1 interstitial duplication (which contains *KDM4C*) found in the proband. While the variant was not present in the mother, paternal inheritance cannot be ruled in or out; however, the proband’s clinical presentation aligns more with the phenotype seen in the Bedouin family, rather than *KDM4C-*associated disorders such as schizophrenia and autism (Kato et al. 2020). Furthermore, it is also generally accepted that genome duplications tend to be better tolerated than deletions (Brewer et al. 1999).

Our population analysis of *SLC30A9* variants in gnomAD also has several limitations. While the gnomAD database attempts to exclude individuals with severe pediatric disease and their first-degree relatives, it is still possible for some individuals with severe disease to be included in the dataset. Furthermore, given that the allele counts are small, the power of the analysis was limited. It is also important to note that individuals with African ancestry are underrepresented in genomics research and may therefore erroneously appear to have a smaller risk of genetic disease (Bentley, Callier, and Rotimi 2020).

## Data Availability

The data that support the findings of this study are available on request from the corresponding author. The data are not publicly available due to privacy or ethical restrictions.

