## Supplementary Information for "Autosomal recessive *SLC30A9* Mutations in a Proband with a Cerebro-Renal Syndrome and No Parental Consanguinity"

**Additional Information on *SLC30A9***

Although the Perez et al. reported that the subcellular localization of the protein is unaffected by the mutation, it is important to note that its localization in wildtype cells has not yet been well characterized. Confocal analysis using *SLC30A9* fused to enhanced green fluorescent protein (EGFP) in a neuroblastoma cell line found that the protein is localized in cytosolic vesicles likely associated with the endoplasmic reticulum. However, fluorescence was not observed in the nucleus, which does not align with previous findings suggesting its role as a nuclear regulator (Perez et al. 2017). It can be speculated that the 63.5 kDa SLC30A9 protein, when fused to the 27 kDa EGFP plasmids, had too low of a nuclear diffusion coefficient to produce a detectable signal (Dross et al. 2009; Wei et al. 2003).

Furthermore, a preprint posted in April 2021 suggests that SLC30A9 acts as a mitochondrial zinc exporter. After exposure to high zinc concentrations, it was reported that the mitochondria in SLC30A9 knockdown HeLa cells had substantially higher zinc concentrations (as measured by the divalent-cation sensitive Rhod-2,AM fluorescent dye) when compared to those in control HeLa cells. The authors also stated that cell toxicity occurs when the SLC30A9 nuclear localization signal is deleted (Kowalczyk et al. 2021). Given that mitochondria have previously been recognized as a possible free zinc storage site, these findings further highlight the importance of SLC30A9 in zinc homeostasis (Lu et al. 2016).

**Clinical Presentation:**

The proband was born at full term gestation via an uncomplicated vaginal delivery to a mother (age range 20 - 25) (**Figure 1**). There was no notable maternal family history of consanguinity, intellectual/developmental disabilities, (I/DD), or seizures. The father reportedly had a relative with developmental delay. The mother reported having regular prenatal visits and ultrasound examinations, during which a two-vessel umbilical cord was noted. The fetus was reportedly active, and the pregnancy was otherwise unremarkable. The mother denied medication use, pre-eclampsia, illness, or chemical exposure during pregnancy.

At birth, the proband was jaundiced and small for gestational age, weighing 5 lbs (2.3 kg) at birth. She required ventilator support but was discharged two days after birth. The mother reported that an abdominal ultrasound was performed on the infant prior to discharge which revealed no abnormalities.

An evaluation by a pediatric neurologist at around age range 0.5 – 1 years noted truncal hypotonia, slow weight gain, and severe global developmental delay. The proband smiled, laughed responsively and cooed, but did not babble. She was able to fixate on, and track objects. No developmental regression was noted. The head circumference was below the second percentile for her age (37.5 cm), and the height and weight were below the third percentiles. A neurological exam was otherwise unremarkable, with normal hearing noted. A motor exam revealed a normal appendicular tone, good head control, and no growth arrest.

As per medical records, neuro magnetic resonance imaging (MRI) at around age range 0.5 – 2 years revealed possible microcephaly, bilateral white matter loss in the frontal and parietal lobes, pachygyria, agenesis of the corpus callosum, and an arachnoid cyst. There was no evidence of hydrocephalus, heterotopic gray matter, mass effect, or midline shift. Evaluation of the ventricles, myelination patterns, brainstem, cerebellum, pituitary glands, and vascular flow voids was unremarkable. A skull X-ray conducted several days later revealed patent coronal, sagittal, and lambdoid sutures indicating no evidence of craniosynostosis. Given that the primary data for the MRI and X-rays are not available, imaging is not shown this report.

This patient first presented to our clinic and agency for services evaluation from the Office of People with Developmental Disabilities in New York State around age range 0.5 - 2 years, after referral for microcephaly and developmental delay. The proband was able to sit without support but would bend forward while doing so. While standing, she was unable to bear weight. The proband smiled socially but did not vocalize or laugh. She was receiving speech, physical, and occupational therapy with little improvement. Feeding difficulties were also noted.

Physical examination revealed a height, weight, and head circumference at or below the third percentile (70.5 cm, 7.7 kg, and 40 cm respectively), which is consistent with previous evaluations. Hypotonia was noted in the trunk and face. Other observed craniofacial abnormalities included arching eyebrows, up-slanting palpebral fissures, and long eyelashes. The parents did not consent for publication of facial photographs. No organomegaly, heart murmurs, or neurological abnormalities were observed. However, it was noted that the proband did not respond to sound. She was subsequently scheduled for an auditory brainstem response (ABR) and hearing evaluation which revealed bilateral sensorineural hearing loss.

A follow-up plasma acylcarnitine profile was not specific for a metabolic disorder, but was notable for mildly elevated glutarylcarnitine, malonylcarnitine, decanoylcarnitine, and 3-hydroxy-tetradecenoyl carnitine levels. Further blood work revealed an elevated serum creatine kinase-MB.

Cytogenetic analysis of phytohaemagglutinin-stimulated cell cultures revealed a normal female karyotype (46,XX) with unremarkable GTG banding patterns. A whole genome single nucleotide polymorphism (SNP) chromosomal microarray conducted using the Affymetrix Cytoscan HD platform and the Chromosome Analysis Suite revealed a 505 kb interstitial duplication of 9p24.1-p24.1. This was investigated as a familial variant, but a fluorescence in situ hybridization (FISH) analysis of maternal blood did not suggest that the variant was maternally inherited. Unfortunately, a paternal blood sample was unable to be obtained for FISH analysis. The identified copy number variant (CNV) in chromosomal region 9p24-p23 includes *KDM4C*, which encodes a lysine demethylase that has been previously implicated in the development and/or progression of certain cancers such as esophageal squamous cell carcinoma (Yang et al. 2000). Consequently, it was considered to be a variant of unknown significance. However, a recent analysis of a Japanese sample set found significant associations between CNVs involving *KDM4C* and neuropsychiatric disorders such as schizophrenia and autism spectrum disorder (Kato et al. 2020).

Routine blood work at around age range 1-4 years revealed elevated blood urea nitrogen (BUN) and serum creatinine levels. The patient was subsequently referred to a pediatric nephrologist. Additional work-up confirmed the above laboratory findings and also revealed lymphocytopenia, elevated cystatin C levels, and elevated parathyroid hormone (PTH) levels. The elevated PTH was suggestive of secondary hyperparathyroidism. The proband was prescribed daily calcitriol supplementation, although prior vitamin D levels were within normal limits. The glomerular filtration rate (GFR) was considered "poor," and estimated to be between 25 and 50 mL/minute. She was referred for a renal ultrasound examination, which revealed relatively small kidneys (5 cm each), but normal echotexture. Hydronephrosis was not noted. She was subsequently diagnosed with Stage 3 chronic kidney disease (CKD).

The patient's next follow-up visit was around age range 5-10 years. She was developmentally delayed, non- ambulatory and non- verbal. Due to food aversion and not eating a sufficient amount of food, she had a gastrostomy tube inserted, although the time of placement was unclear. Physical exam was notable for microcephaly, dysmorphic facial features, and hypotonia. New manifestations including constant facial mimicking and dystonic arm movements. She continued to be followed by a pediatric nephrologist, who reported a "relatively stable" GFR of 40-50 ml/min.  She had global developmental delay. It was also documented that she also received cochlear implants several years prior to this, and was reportedly able to hear and occasionally respond to commands. At another visit two months later, physical exam revealed no improvement in her microcephaly and motor development and was now notable for constant dystonia affecting the entire body. Blood samples were collected from the mother, father, and the proband for research-based whole exome sequencing (WES), but the family was subsequently lost to follow-up. Laboratory analysis of this sample at age seven revealed elevated BUN (38 mg/dL) and creatinine (1.03 mg/dL) levels.

**WES Analysis:**

Blood samples collected from the proband, mother, and father were sent for clinical WES (Novogene, Sacramento, CA). DNA extracted from whole blood was mechanically sheared and prepared as libraries with dual-indexed sequencing barcodes. The SureSelect Human AII Exon V6 capture baits (Agilent Technologies, Santa Clara, CA) were used to enrich the pre-capture libraries. The enriched, post-capture libraries were then sequenced using the NovaSeq 6000 instrument (Illumina, San Diego, CA). Variant calling and interpretation were conducted by the UK state registered clinical scientists using the Congenica platform (Congenica Limited, Cambridge, UK).  Raw WES data was uploaded to the Congenica platform (Congenica Limited, Cambridge, UK) for analysis by UK state registered clinical scientists. The raw FASTQ files were aligned to the GRCh37 reference chromosome using BWA-MEM (0.7.12) and variant calling performed using GATK HaplotypeCaller (version 3.4-46). Quality control (QC) of FASTQ and VCF files were performed using FASTQC (0.11.5) and VariantRecallibrator (GATK) respectively. The Ensembl Variant Effect Predictor (VEP; version 81) was used to annotate VCF files with current RefSeq and Ensembl transcripts. Coverage and mapping statistics for whole exome sequencing performed on the proband, mother, and father are shown in **Supplementary Table 1**.

**Variant Interpretation & Validation:**

Guidelines established by the *UK Association for Clinical Genetic Science* and the American College of Medical Genetics and Genomics were used to guide variant interpretation (Richards et al. 2015; Wallis et al. 2013). Variants were first prioritized based on the Exomiser's Gene Pheno Score (version 10), and then filtered if their allele frequency was greater than 0.5% in EXAC, NHLBI Exome Sequencing Project (ESP), UK10K and 1000 genomes (Tennessen et al. 2012; 1000 Genomes Project Consortium et al. 2015). Although the c.40delA variant has been reported in Genome Aggregation Database (gnomAD), it has been flagged as having "dubious" annotation or quality (Karczewski et al. 2020). The resulting variant list was further refined through the use of variant consequence filters and inheritance filtering; only variants predicted to impact coding sequences, consensus splice sites, and splice regions were considered to be variants of interest and included for further study. Two distinct, likely pathogenic, compound heterozygous candidate variants in *SLC30A9* were detected in the proband (NM_006345 c.40delA and c.86_87dupCC), consistent with autosomal-recessive inheritance (**Supplementary Table 2**). Both variants are predicted to result in nonsense mediated decay and concomitant loss of the encoded protein. Combined Annotation Dependent Depletion (CADD) scores were also calculated for variants (Rentzsch et al. 2019). The following American College of Medical Genetics and Genomics (ACMG) evidence criteria suggesting pathogenicity have been met: frameshift (i.e., null) variants in a gene where loss of function mutations are a known mechanism of disease (PVS1), well-established *in vitro* studies demonstrating a damaging effect on the gene product, detected in *trans* as a pathogenic variant (PM3), and proband's phenotype is highly specific for the gene (PP4). These criteria and other bioinformatic results are also shown in **Supplementary Table 2.**

**Sanger Sequencing:**

Direct sequencing was performed on an ABI3130 automated sequencer using version 1.1 of the Big Dye fluorescent method according to the manufacturer's instructions (Thermo Fisher Scientific, MA). Sequence data were analyzed using the Sequencher program (GeneCode Corp, MI). The two sets of primers used for targeted sequencing of the two variants within the *SLC30A9* gene (c.40delA and c.86_87dupCC, NM_006345) are respectively:

Genomics and the Association for Molecular Pathology." *Genetics in Medicine: Official Journal of the American College of Medical Genetics* 17 (5): 405–24.

Tennessen, Jacob A., Abigail W. Bigham, Timothy D. O'Connor, Wenqing Fu, Eimear E. Kenny, Simon Gravel, Sean McGee, et al. 2012. "Evolution and Functional Impact of Rare Coding Variation from Deep Sequencing of Human Exomes." *Science* 337 (6090): 64–69.

Wallis, Yvonne, Stewart Payne, Ciaron McAnulty, Danielle Bodmer, Erik Sistermans, Kathryn Robertson, David Moore, Stephen Abbs, Zandra Deans, and Andrew Devereau. 2013. "Practice Guidelines for the Evaluation of Pathogenicity and the Reporting of Sequence Variants in Clinical Molecular Genetics." *Association for Clinical Genetic Science and the Dutch Society of Clinical Genetic Laboratory Specialists*. http://dx.doi.org/.

Wei, Xunbin, Vanessa G. Henke, Carsten Strübing, Edward B. Brown, and David E. Clapham. 2003. "Real-Time Imaging of Nuclear Permeation by EGFP in Single Intact Cells." *Biophysical Journal* 84 (2 Pt 1): 1317–27.

Yang, Z. Q., I. Imoto, Y. Fukuda, A. Pimkhaokham, Y. Shimada, M. Imamura, S. Sugano, Y. Nakamura, and J. Inazawa. 2000. "Identification of a Novel Gene, GASC1, within an Amplicon at 9p23-24 Frequently Detected in Esophageal Cancer Cell Lines." *Cancer Research* 60 (17): 4735–39.

SLC30A9_40F: 5'-GGTC4CTGAGGAGAATGTATTG-3'

SLC30A9_40R: 5'-GCCAGAAAGGAAGGTTCTGA-3'22000.0

SLC30A9_86F: 5'- CAACCAAGCGCAAAAGTACA-3'

SLC30A9_86R: 5'- TTTGGTACAAAGCGGTAGGC-3'

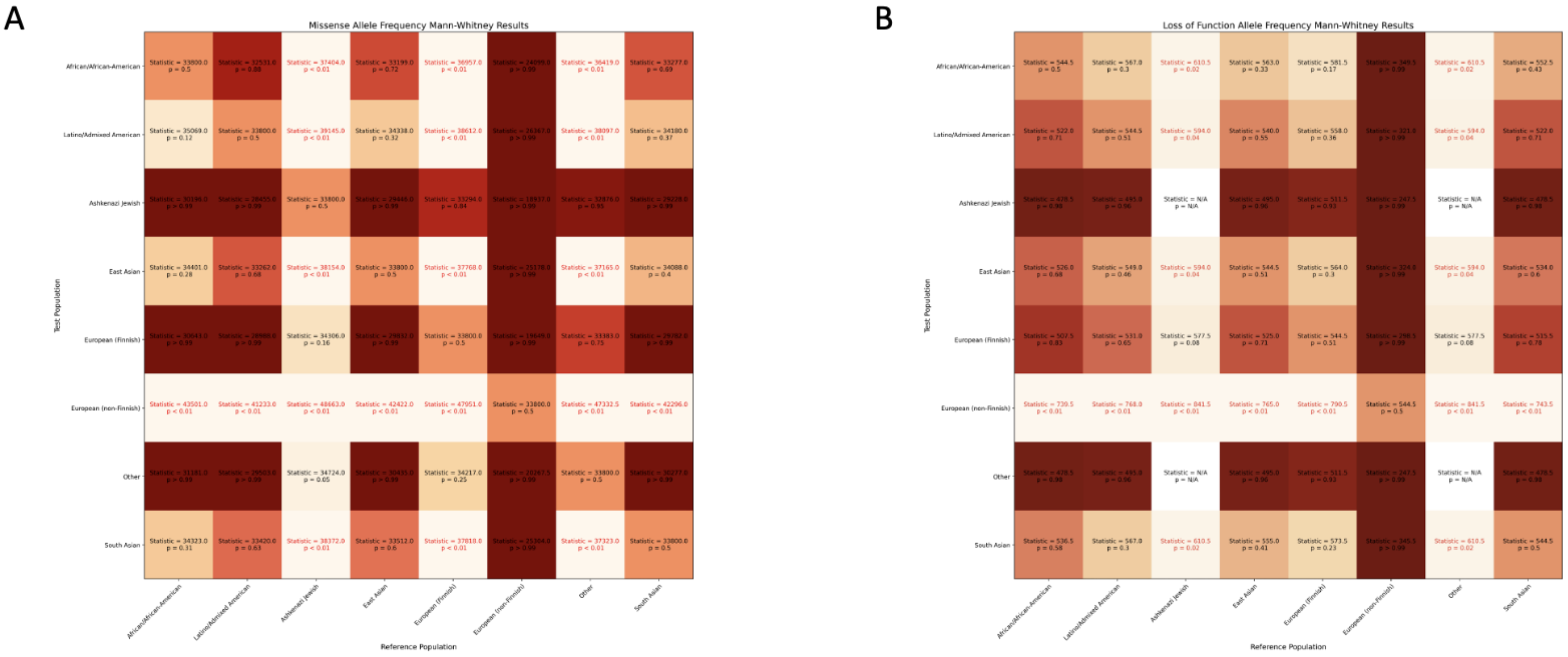


Supplementary Figure 1: Heatmaps visualizing the results of one-sided Mann-Whitney Tests of significance for allele frequencies (AFs) of *SLC30A9* variants between populations in the Genome Aggregation Database (gnomAD). The alternate hypothesis tested is that the AF of the "Test" population (left) is greater than the "Reference" population (bottom). A lighter fill color denotes a lower p-value, while darker colors denote a higher p-value. Boxes with red text signify that the p-value was less than 0.05. A) depicts the results for missense variants, B) shows the results for loss of function variants.

Supplementary Table 1: Coverage and mapping statistics for whole exome sequencing performed on the proband, mother, and father.

| | Proband | Mother | Father |
|---|---|---|---|
| Average Read Depth at Target Regions | 159 | 120 | 125 |
| Mismatch Rate at Target Regions | 0.30% | 0.31% | 0.35% |

Supplementary Table 2: *SLC30A9* variants identified in the proband, as well as their protein consequence, predicted effect, parent of origin, and Combined Annotation Dependent Depletion (CADD) score.

| Gene | Chromosome | Position (hg19) | HGVS DNA Reference | HGVS Protein Reference | Variant Type | Predicted Effect | dbSNP/dbVar ID | Genotype | ClinVar ID | Parent of Origin | CADD Score | HGVS Evidence Strength |
|---|---|---|---|---|---|---|---|---|---|---|---|---|
| *SLC30A9* | 4 | 41,992,707 | NM_006345.3: c.40delA | p.S14AfsX28 | Deletion | Frame-shift | rs767078182 | Heterozygous | In process | Paternal | 23.8 | PVS1 PS3 PM3 PP4 |
| *SLC30A9* | 4 | 41,992,753 | NM_006345.3: c.86_87dupCC | p.C30PfsX13 | Duplication | Frame-shift | rs752245649 | Heterozygous | In process | Maternal | N/A | |

Supplementary Table 3: Allele frequency (AF) summary statistics by population for reported missense and loss of function variants in individuals unaffected by severe pediatric disease. Data was accessed in gnomAD on March 14, 2021.

| Population | Number of Variants Observed in Population / Total Number of Variants Observed | Mean AF | Standard Deviation of AF | Minimum AF | 25th Percentile AF | Median AF | 75th Percentile AF | Maximum AF |
|---|---|---|---|---|---|---|---|---|
| **Missense Variants** | | | | | | | | |
| African/African-American | 34/260 | 2.81E-05 | 0.00017079 | 0 | 0 | 0 | 0 | 0.00250283 |
| Latino/Admixed American | 48/260 | 1.70E-05 | 0.00010946 | 0 | 0 | 0 | 0 | 0.00167022 |
| Ashkenazi Jewish | 6/260 | 1.36E-05 | 0.00013852 | 0 | 0 | 0 | 0 | 0.0017445 |

| | | | | | | | | |
|---|---|---|---|---|---|---|---|---|
| | | | | | | | | 2 |
| East Asian | 40/260 | 2.44E-05 | 9.61E-05 | 0 | 0 | 0 | 0 | 0.0006426 7 |
| European (Finnish) | 10/260 | 6.49E-06 | 5.98E-05 | 0 | 0 | 0 | 0 | 0.0009179 4 |
| European (non- Finnish) | 123/260 | 1.96E-05 | 0.00016125 | 0 | 0 | 0 | 8.93E-06 | 0.0025843 2 |
| Other | 13/260 | 1.34E-05 | 0.0001083 | 0 | 0 | 0 | 0 | 0.0016759 8 |
| South Asian | 42/260 | 2.73E-05 | 0.00023539 | 0 | 0 | 0 | 0 | 0.0029052 5 |
| Loss of Function Variants | | | | | | | | |
| African/ African-American | 4/33 | 1.75E-05 | 5.09E-05 | 0 | 0 | 0 | 0 | 0.00020850 70892 |
| tino/Admixed Americ | 3/33 | 2.62E-06 | 8.42E-06 | 0 | 0 | 0 | 0 | 2.92E-05 |

| | | | | | | | | |
|---|---|---|---|---|---|---|---|---|
| Ashkenazi Jewish | 0/33 | 0.00E+00 | 0 | 0 | 0 | 0 | 0 | 0 |
| East Asian | 3/33 | 2.28E-05 | 1.12E-04 | 0 | 0 | 0 | 0 | 0.0006418485237 |
| European (Finnish) | 2/33 | 2.80E-06 | 1.12E-05 | 0 | 0 | 0 | 0 | 4.63E-05 |
| European (non-Finnish | 18/33 | 6.93E-06 | 8.31E-06 | 0 | 0 | 8.81E-06 | 8.99E-06 | 3.52E-05 |
| Other | 0/33 | 0.00E+00 | 0 | 0 | 0 | 0 | 0 | 0 |
| South Asian | 4/33 | 4.01E-06 | 1.10E-05 | 0 | 0 | 0 | 0 | 3.38E-05 |
