## Supplemental File 1 for "Autosomal recessive *SLC30A9* Mutations in a Proband with a Cerebro-Renal Syndrome and No Parental Consanguinity"

### Population Analysis of *SLC30A9* Variants in gnomAD

Karczewski, K.J., Francioli, L.C., Tiao, G. et al. The mutational constraint spectrum quantified from variation in 141,456 humans. Nature 581, 434–443 (2020). https://doi.org/10.1038/s41586-020-2308-7

#### Analysis Setup – Import of Libraries & Data

In [1]:
```
import pandas as pd
from scipy.stats import normaltest, kruskal, mannwhitneyu
import numpy as np
import matplotlib.pyplot as plt
import math
```

In [2]:
```
# Importing gnomAD Data (As of 03/14/2021)
all_missense = pd.read_csv('gnomAD_v2.1.1_ENSG00000014824_2021_03_14_13_53_29
lof = pd.read_csv('gnomAD_v2.1.1_ENSG00000014824_2021_03_14_13_53_34.csv')
```

In [3]:
```
all_missense.columns
```

Out[3]:
```
Index(['Chromosome', 'Position', 'rsID', 'Reference', 'Alternate', 'Source',
       'Filters - exomes', 'Filters - genomes', 'HGVS Consequence',
       'Protein Consequence', 'Transcript Consequence', 'VEP Annotation',
       'ClinVar Clinical Significance', 'Flags', 'Allele Count',
       'Allele Number', 'Allele Frequency', 'Homozygote Count',
       'Hemizygote Count', 'Allele Count African/African-American',
       'Allele Number African/African-American',
       'Homozygote Count African/African-American',
       'Hemizygote Count African/African-American',
       'Allele Count Latino/Admixed American',
       'Allele Number Latino/Admixed American',
       'Homozygote Count Latino/Admixed American',
       'Hemizygote Count Latino/Admixed American',
       'Allele Count Ashkenazi Jewish', 'Allele Number Ashkenazi Jewish',
       'Homozygote Count Ashkenazi Jewish',
       'Hemizygote Count Ashkenazi Jewish', 'Allele Count East Asian',
       'Allele Number East Asian', 'Homozygote Count East Asian',
       'Hemizygote Count East Asian', 'Allele Count European (Finnish)',
       'Allele Number European (Finnish)',
       'Homozygote Count European (Finnish)',
       'Hemizygote Count European (Finnish)',
       'Allele Count European (non-Finnish)',
       'Allele Number European (non-Finnish)',
       'Homozygote Count European (non-Finnish)',
       'Hemizygote Count European (non-Finnish)', 'Allele Count Other',
       'Allele Number Other', 'Homozygote Count Other',
       'Hemizygote Count Other', 'Allele Count South Asian',
       'Allele Number South Asian', 'Homozygote Count South Asian',
       'Hemizygote Count South Asian'],
      dtype='object')
```

In [4]:
```
all_missense['rsID'].to_clipboard(index=False)
```

In [5]:
```
all_missense[['Chromosome', 'Position', 'rsID', 'Reference', 'Alternate']].to
```

In [6]:
```
# Removing variants with a homozygote count greater than zero
all_missense = all_missense.loc[all_missense['Homozygote Count']==0]
lof = lof.loc[lof['Homozygote Count']==0]
```

In [7]:
```
# Displaying columns in dataset
if all(all_missense.columns == lof.columns):
    print(all_missense.columns)
```

```
Index(['Chromosome', 'Position', 'rsID', 'Reference', 'Alternate', 'Source',
       'Filters - exomes', 'Filters - genomes', 'HGVS Consequence',
       'Protein Consequence', 'Transcript Consequence', 'VEP Annotation',
       'ClinVar Clinical Significance', 'Flags', 'Allele Count',
       'Allele Number', 'Allele Frequency', 'Homozygote Count',
       'Hemizygote Count', 'Allele Count African/African-American',
       'Allele Number African/African-American',
       'Homozygote Count African/African-American',
       'Hemizygote Count African/African-American',
       'Allele Count Latino/Admixed American',
       'Allele Number Latino/Admixed American',
       'Homozygote Count Latino/Admixed American',
       'Hemizygote Count Latino/Admixed American',
       'Allele Count Ashkenazi Jewish', 'Allele Number Ashkenazi Jewish',
       'Homozygote Count Ashkenazi Jewish',
       'Hemizygote Count Ashkenazi Jewish', 'Allele Count East Asian',
       'Allele Number East Asian', 'Homozygote Count East Asian',
       'Hemizygote Count East Asian', 'Allele Count European (Finnish)',
       'Allele Number European (Finnish)',
       'Homozygote Count European (Finnish)',
       'Hemizygote Count European (Finnish)',
       'Allele Count European (non-Finnish)',
       'Allele Number European (non-Finnish)',
       'Homozygote Count European (non-Finnish)',
       'Hemizygote Count European (non-Finnish)', 'Allele Count Other',
       'Allele Number Other', 'Homozygote Count Other',
       'Hemizygote Count Other', 'Allele Count South Asian',
       'Allele Number South Asian', 'Homozygote Count South Asian',
       'Hemizygote Count South Asian'],
      dtype='object')
```

In [8]:
```
all_missense.head()
```

Out[8]:

| | Chromosome | Position | rsID | Reference | Alternate | Source | Filters - exomes | Filters - genomes |
|---|---|---|---|---|---|---|---|---|
| **0** | 4 | 41992673 | rs1185749484 | T | C | gnomAD Exomes | PASS | NaN |
| **1** | 4 | 41992676 | rs767957901 | C | G | gnomAD Exomes | PASS | NaN |
| **2** | 4 | 41992676 | rs767957901 | C | T | gnomAD Exomes | PASS | NaN |
| **3** | 4 | 41992682 | rs754541096 | T | TGGC | gnomAD Exomes | PASS | NaN |
| **4** | 4 | 41992684 | rs1385920478 | G | T | gnomAD Exomes | PASS | NaN |

5 rows × 51 columns

In [9]:
```
lof.head()
```

Out[9]:

| | Chromosome | Position | rsID | Reference | Alternate | Source | Filters - exomes | Filt geno |
|---|---|---|---|---|---|---|---|---|
| **0** | 4 | 41992707 | rs767078182 | TA | T | gnomAD Exomes,gnomAD Genomes | PASS | [illegible] |
| **1** | 4 | 41992712 | rs769918635 | G | A | gnomAD Exomes | PASS | |
| **2** | 4 | 41992753 | rs752245649 | G | GCC | gnomAD Exomes,gnomAD Genomes | PASS | [illegible] |
| **3** | 4 | 42003631 | rs776947278 | A | T | gnomAD Exomes | PASS | |
| **4** | 4 | 42003666 | rs751753033 | C | A | gnomAD Exomes | PASS | |

5 rows × 51 columns

In [10]:
```
# List of all population categories in gnomAD
gnomad_ethnicities = ['African/African-American', 'Latino/Admixed American',
                      'East Asian', 'European (Finnish)', 'European (non-Finnish)',
```

#### Analysis of Missense Variants

In [11]:

```
# Selecting only total allele frequencies
all_missense_total_af = all_missense['Allele Frequency']

# Creating a dictionary with computed allele frequencies for each population
missense_af_by_ethnicity = {}
for ethnicity in gnomad_ethnicities:
    missense_af_by_ethnicity[ethnicity] = all_missense['Allele Count' + ' '+ [illegible]
    all_missense['Allele Number' + ' '+ ethnicity]
    missense_af_by_ethnicity[ethnicity] = missense_af_by_ethnicity[ethnicity]
```

In [12]:

```
for k in missense_af_by_ethnicity.keys():
    print(k, sum(missense_af_by_ethnicity[k]>0))
```

```
African/African-American 34
Latino/Admixed American 48
Ashkenazi Jewish 6
East Asian 40
European (Finnish) 10
European (non-Finnish) 123
Other 13
South Asian 42
```

In [13]:

```
# Showing summary Statistics
pd.DataFrame(missense_af_by_ethnicity).describe()
```

Out[13]:

| | African/African-American | Latino/Admixed American | Ashkenazi Jewish | East Asian | European (Finnish) | European (non-Finnish) | |
|---|---|---|---|---|---|---|---|
| **count** | 260.000000 | 260.000000 | 260.000000 | 260.000000 | 260.000000 | 260.000000 | 26 |
| **mean** | 0.000028 | 0.000017 | 0.000014 | 0.000024 | 0.000006 | 0.000020 | |
| **std** | 0.000171 | 0.000109 | 0.000139 | 0.000096 | 0.000060 | 0.000161 | |
| **min** | 0.000000 | 0.000000 | 0.000000 | 0.000000 | 0.000000 | 0.000000 | |
| **25%** | 0.000000 | 0.000000 | 0.000000 | 0.000000 | 0.000000 | 0.000000 | |
| **50%** | 0.000000 | 0.000000 | 0.000000 | 0.000000 | 0.000000 | 0.000000 | |
| **75%** | 0.000000 | 0.000000 | 0.000000 | 0.000000 | 0.000000 | 0.000009 | |
| **max** | 0.002503 | 0.001670 | 0.001745 | 0.000643 | 0.000918 | 0.002584 | |

In [14]:

```
# Displaying a bar graph with mean allele frequency by ethnicity
plt.figure(num=None, figsize=(20, 6), dpi=80, facecolor='w', edgecolor='k')
plt.bar(missense_af_by_ethnicity.keys(), [np.mean(v) for v in missense_af_by_[illegible]
        color='g')
plt.show()
```

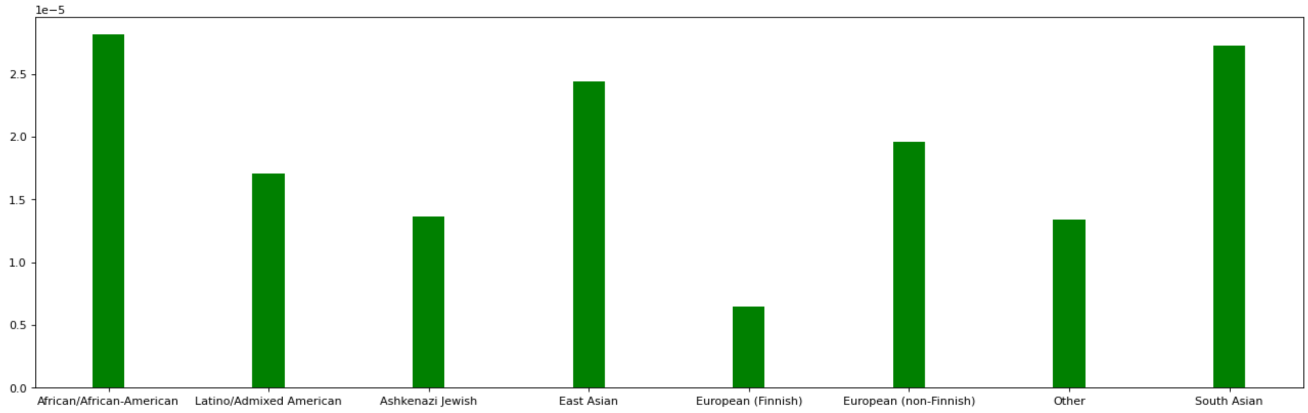


#### Performing the D'Agostino-Pearson Test for Normality:

```
Ho: The data is normally distributed.
HA: The data is NOT normally distributed.
α = 0.05
```

In [15]:
```
normaltest(all_missense_total_af) # Suggests that the data is NOT normally di
```

Out[15]:
```
NormaltestResult(statistic=571.6015762955817, pvalue=7.556049316854983e-125)
```

Given that the p-value is less than 0.05, Ho is rejected and it is concluded that the data is unlikely to be normally distributed. As a result, parametric tests such as the Student's T-Test and the One-Way Analysis of Variance (ANOVA) cannot be used.

#### Performing a Kruskal-Wallis One-Way ANOVA

```
Ho: There are NO differences in allele frequencies between
populations.
HA: There IS A difference in allele frequencies between
populations.
α = 0.05
```

In [16]:
```
# Performing a Kruskal-Wallis One-Way Analysis of Variance
kruskal(missense_af_by_ethnicity.get('African/African-American'),
        missense_af_by_ethnicity.get('Latino/Admixed American'),
        #missense_af_by_ethnicity.get('Ashkenazi Jewish'), #
        #missense_af_by_ethnicity.get('East Asian'),
        missense_af_by_ethnicity.get('European (Finnish)'),
        #missense_af_by_ethnicity.get('European (non-Finnish)'), #
        #missense_af_by_ethnicity.get('Other'), #
       missense_af_by_ethnicity.get('South Asian')
       )
```

Out[16]:
```
KruskalResult(statistic=25.740247428089855, pvalue=1.0809515958409424e-05)
```

Given that the p-value is less than 0.05, Ho is rejected and it is concluded that there is a statistical evidence to suggest that there is a difference in allele frequencies between populations.

##### Performing a Mann-Whitney U Test/Wilcoxon Rank-Sum Test Between the African/African-American Population & Other Populations

A one-sided test will be used, with the following null and alternate hypothesis:

```
Ho: The allele frequencies in the African/African-American
population are NOT greater than in the other
    population.
HA: The allele frequencies in the African/African-American
population are NOT greater than in the other
    population.
α = 0.05
```

In [17]:

```
for ethnicity in gnomad_ethnicities:
    if ethnicity != 'African/African-American':
        print(ethnicity + ': ' + str(mannwhitneyu(missense_af_by_ethnicity.ge
                                  missense_af_by_ethnicity.get(ethnicity),
                                  alternative='greater')))
```

```
Latino/Admixed American: MannwhitneyuResult(statistic=32531.0, pvalue=0.878629
3258404441)
Ashkenazi Jewish: MannwhitneyuResult(statistic=37404.0, pvalue=2.6504425979546
166e-06)
East Asian: MannwhitneyuResult(statistic=33199.0, pvalue=0.7183514500175109)
European (Finnish): MannwhitneyuResult(statistic=36957.0, pvalue=6.74718238012
941e-05)
European (non-Finnish): MannwhitneyuResult(statistic=24099.0, pvalue=0.9999999
999984318)
Other: MannwhitneyuResult(statistic=36419.0, pvalue=0.001059536642508388)
South Asian: MannwhitneyuResult(statistic=33277.0, pvalue=0.690526540621231)
```

The results suggest the African/African-American population may have a higher allele frequency of mutations than the *Ashkenazi Jewish*, and *Other* populations. However, it also appears that the *European (non-Finnish)* population may have a significantly higher allele frequency. This is tested below:

A one-sided test will be used, with the following null and alternate hypothesis:

```
Ho: The allele frequencies in the European (non-Finnish)
population are NOT greater than in the other
    population.
HA: The allele frequencies in the European (non-Finnish)
population are NOT greater than in the other
    population.
α = 0.05
```

In [18]:

```
for ethnicity in gnomad_ethnicities:
    if ethnicity != 'European (non-Finnish)':
        print(ethnicity + ': ' + str(mannwhitneyu(missense_af_by_ethnicity.ge
                                  missense_af_by_ethnicity.get(ethnicity),
                                  alternative='greater')))
```

```
African/African-American: MannwhitneyuResult(statistic=43501.0, pvalue=1.57623
30724328924e-12)
Latino/Admixed American: MannwhitneyuResult(statistic=41233.0, pvalue=1.028928
2197046232e-07)
Ashkenazi Jewish: MannwhitneyuResult(statistic=48663.0, pvalue=1.2883472029226
573e-30)
East Asian: MannwhitneyuResult(statistic=42422.0, pvalue=4.710077041305666e-10
)
European (Finnish): MannwhitneyuResult(statistic=47951.0, pvalue=2.29389493704
96507e-27)
Other: MannwhitneyuResult(statistic=47332.5, pvalue=8.0405128846415705e-25)
South Asian: MannwhitneyuResult(statistic=42296.0, pvalue=9.534115212619336e-1
0)
```

Given that $p < 0.05$ for all tests run, it is concluded that there is statistically significant evidence to suggest that the allele frequencies are higher in the *European (non-Finnish)* than in any of the other populations studied.

#### Analysis of Loss of Function Variants

In [19]:

```
# Selecting only total allele frequencies
LOF_total_af = lof['Allele Frequency']

# Creating a dictionary with computed allele frequencies for each population
lof_af_by_ethnicity = {}
for ethnicity in gnomad_ethnicities:
    lof_af_by_ethnicity[ethnicity] = lof['Allele Count' + ' '+ ethnicity] / \
    lof['Allele Number' + ' '+ ethnicity]
    lof_af_by_ethnicity[ethnicity] = lof_af_by_ethnicity[ethnicity].fillna(0)
```

In [20]:

```
for k in lof_af_by_ethnicity.keys():
    print(k, sum(lof_af_by_ethnicity[k]>0))
```

```
African/African-American 4
Latino/Admixed American 3
Ashkenazi Jewish 0
East Asian 3
European (Finnish) 2
European (non-Finnish) 18
Other 0
South Asian 4
```

In [21]:

```
# Showing summary Statistics
pd.DataFrame(lof_af_by_ethnicity).describe().to_clipboard()
```

In [22]:

```
# Displaying a bar graph with mean allele frequency by ethnicity
plt.figure(num=None, figsize=(20, 6), dpi=80, facecolor='w', edgecolor='k')
plt.bar(lof_af_by_ethnicity.keys(), [np.mean(v) for v in lof_af_by_ethnicity.
        color='g')
plt.show()
```

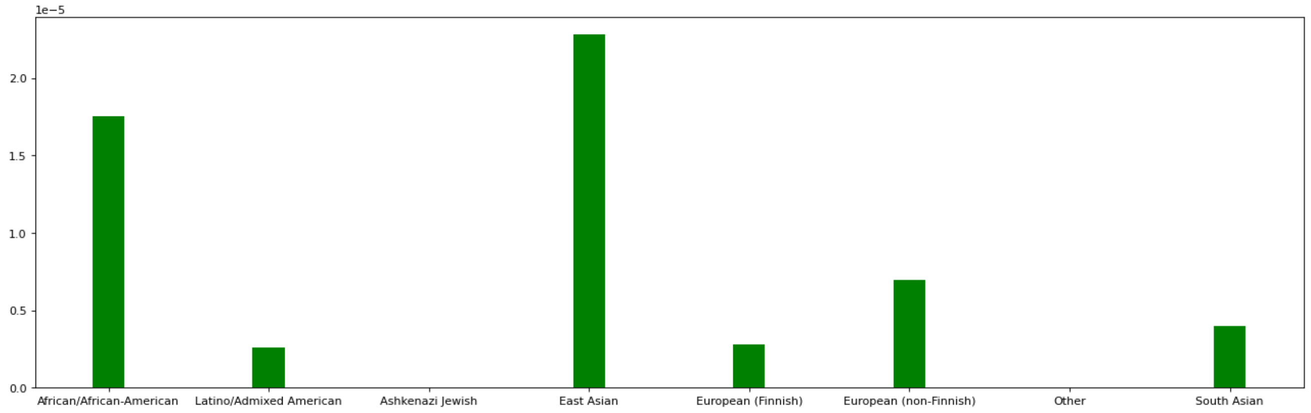


##### Performing the D’Agostino-Pearson Test for Normality:

```
Ho: The data is normally distributed.
HA: The data is NOT normally distributed.
α = 0.05
```

In [23]:
```
normaltest(LOF_total_af) # Suggests that the data is NOT normally distributed
```

Out[23]: `NormaltestResult(statistic=30.790751813467693, pvalue=2.0600283487264053e-07)`

Given that the p-value is less than 0.05, Ho is rejected and it is concluded that the data is unlikely to be normally distributed. As a result, parametric tests such as the Student's T-Test and the One-Way Analysis of Variance (ANOVA) cannot be used.

##### Performing a Kruskal-Wallis One-Way ANOVA

```
Ho: There are NO differences in allele frequencies between
populations.
HA: There IS A difference in allele frequencies between
populations.
α = 0.05
```

In [24]:
```
# Performing a Kruskal-Wallis One-Way Analysis of Variance
kruskal(lof_af_by_ethnicity.get('African/African-American'),
        lof_af_by_ethnicity.get('Latino/Admixed American'),
        lof_af_by_ethnicity.get('Ashkenazi Jewish'), #
        lof_af_by_ethnicity.get('East Asian'),
        lof_af_by_ethnicity.get('European (Finnish)'),
        lof_af_by_ethnicity.get('European (non-Finnish)'), #
        lof_af_by_ethnicity.get('Other'), #
       lof_af_by_ethnicity.get('South Asian')
       )
```

Out[24]: `KruskalResult(statistic=54.708922629582645, pvalue=1.702585808979608e-09)`

Given that the p-value is less than 0.05, Ho is rejected and it is concluded that there is a statistical evidence to suggest that there is a difference in allele frequencies between populations.

##### Performing a Mann-Whitney U Test/Wilcoxon Rank-Sum Test Between the African/African-American Population & Other Populations

A one-sided test will be used, with the following null and alternate hypothesis:

```
Ho: The allele frequencies in the African/African-American
population are NOT greater than in the other
    population.
HA: The allele frequencies in the African/African-American
population are NOT greater than in the other
    population.
α = 0.05
```

In [25]:
```
for ethnicity in gnomad_ethnicities:
    if ethnicity != 'African/African-American':
        print(ethnicity + ': ' + str(mannwhitneyu(lof_af_by_ethnicity.get('Af
                                      lof_af_by_ethnicity.get(ethnicity),
                                      alternative='greater')))
```

```
Latino/Admixed American: MannwhitneyuResult(statistic=567.0, pvalue=0.29879351
71673709)
Ashkenazi Jewish: MannwhitneyuResult(statistic=610.5, pvalue=0.021124389952612
073)
East Asian: MannwhitneyuResult(statistic=563.0, pvalue=0.3329082746588814)
European (Finnish): MannwhitneyuResult(statistic=581.5, pvalue=0.1739708540971
8188)
European (non-Finnish): MannwhitneyuResult(statistic=349.5, pvalue=0.998598614
360471)
Other: MannwhitneyuResult(statistic=610.5, pvalue=0.021124389952612073)
South Asian: MannwhitneyuResult(statistic=552.5, pvalue=0.43263543939171734)
```

The results suggest the African/African-American population may have a higher allele frequency of mutations than the *Ashkenazi Jewish*, and *Other* populations. However, it also appears that the *European (non-Finnish)* population may have a significantly higher allele frequency. This is tested below:

A one-sided test will be used, with the following null and alternate hypothesis:

```
Ho: The allele frequencies in the European (non-Finnish)
population are NOT greater than in the other
    population.
HA: The allele frequencies in the European (non-Finnish)
population are NOT greater than in the other
    population.
α = 0.05
```

In [26]:
```
for ethnicity in gnomad_ethnicities:
    if ethnicity != 'European (non-Finnish)':
        print(ethnicity + ': ' + str(mannwhitneyu(lof_af_by_ethnicity.get('Eu
                                      lof_af_by_ethnicity.get(ethnicity),
                                      alternative='greater')))
```

```
African/African-American: MannwhitneyuResult(statistic=739.5, pvalue=0.0014731146588041473)
Latino/Admixed American: MannwhitneyuResult(statistic=768.0, pvalue=0.0002699387709965528)
Ashkenazi Jewish: MannwhitneyuResult(statistic=841.5, pvalue=6.263500577974289e-07)
East Asian: MannwhitneyuResult(statistic=765.0, pvalue=0.00032055380090556534)
European (Finnish): MannwhitneyuResult(statistic=790.5, pvalue=5.420102111955623e-05)
Other: MannwhitneyuResult(statistic=841.5, pvalue=6.263500577974289e-07)
South Asian: MannwhitneyuResult(statistic=743.5, pvalue=0.0012047314835430l6)
```

Given that $p < 0.05$ for all tests run, it is concluded that there is statistically significant evidence to suggest that the allele frequencies are higher in the *European (non-Finnish)* than in any of the other populations studied.

#### Creating Additional Visualizations

##### Missense Variants

In [27]:

```
p_dict = {}
stats_dict = {}
statistics = {}
p_vals = {}
X_list = []
for ethnicity in gnomad_ethnicities:
    statistics = {}
    p_vals = {}
    X_list.append(ethnicity)
    for ethnicity2 in gnomad_ethnicities:
        try:
            X_list.append(ethnicity)
            test_stats = mannwhitneyu(missense_af_by_ethnicity.get(ethnicity2
                                      missense_af_by_ethnicity.get(ethnicity),
                                      alternative='greater')
            statistics[ethnicity2] = test_stats[0]
            p_vals[ethnicity2] = test_stats[1]

        except:
            statistics[ethnicity2] = np.nan
            p_vals[ethnicity2] = np.nan
    p_dict[ethnicity] = p_vals
    stats_dict[ethnicity] = statistics
```

In [28]:

```
p_to_plot = np.transpose(np.array([list(v.values()) for v in p_dict.values()]
stat_to_plot = np.transpose(np.array([list(v.values()) for v in stats_dict.va
X_vars =  np.array([list(v.keys()) for v in p_dict.values()])[0]
y_vars = np.array(list(p_dict.keys()))
```

In [29]:

```python
fig, ax = plt.subplots(figsize=(15,15), dpi=300)
im = ax.imshow(p_to_plot, cmap='OrRd') # cmap='binary_r'
# We want to show all ticks...
ax.set_xticks(np.arange(len(X_vars)))
ax.set_yticks(np.arange(len(y_vars)))
# ... and label them with the respective list entries
ax.set_xticklabels(X_vars)
ax.set_yticklabels(y_vars)

# Rotate the tick labels and set their alignment.
plt.setp(ax.get_xticklabels(), rotation=45, ha="right",
         rotation_mode="anchor")

# Loop over data dimensions and create text annotations.
for i in range(len(y_vars)):
    for j in range(len(X_vars)):
        p_value_text = round(p_to_plot[i, j],2)

        if math.isnan(p_value_text):
            p_value_text = '= N/A'
            text_color_red = False
        elif p_value_text == 1.0:
            p_value_text = '> 0.99'
            text_color_red = False
        elif p_value_text == 0.0:
            p_value_text = '< 0.01'
            text_color_red = True
        else:
            if p_value_text < 0.05:
                text_color_red = True
            else:
                text_color_red = False
            p_value_text = '= {}'.format(p_value_text)

        stat_value_text = round(stat_to_plot[i, j],2)
        if math.isnan(stat_value_text):
            stat_value_text = 'N/A'
        text_to_plot = 'Statistic = {stat}\n p {p}'.format(stat=stat_value_te

        if p_value_text != 'N/A' and text_color_red:
            text = ax.text(j, i, text_to_plot,
                           ha="center", va="center", color="r")
        else:
            text = ax.text(j, i, text_to_plot,
                           ha="center", va="center", color="black")



ax.set_title("Missense Allele Frequency Mann-Whitney Results", size=15)
ax.set_xlabel("Reference Population", size=12)
ax.set_ylabel("Test Population", size=12)
fig.tight_layout()
# plt.show()
fig.savefig('missense_heatmap.svg', format='svg')
```

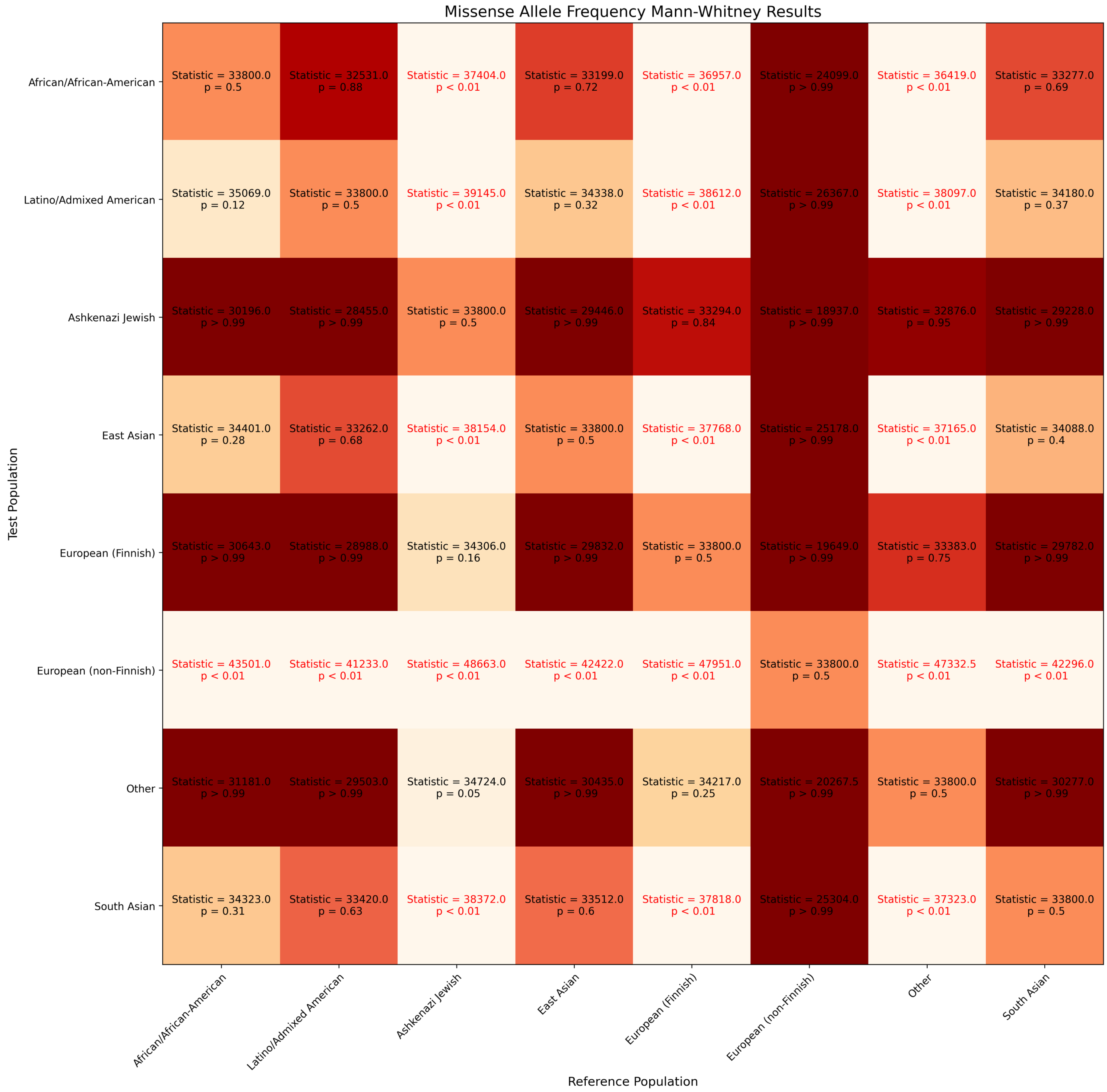


### LoF Variants

In [30]:

```
p_dict = {}
stats_dict = {}
statistics = {}
p_vals = {}
for ethnicity in gnomad_ethnicities:
    statistics = {}
    p_vals = {}
    X_list.append(ethnicity)
    for ethnicity2 in gnomad_ethnicities:
        try:
            X_list.append(ethnicity)
            test_stats = mannwhitneyu(lof_af_by_ethnicity.get(ethnicity2),
                         lof_af_by_ethnicity.get(ethnicity),
                         alternative='greater')
            statistics[ethnicity2] = test_stats[0]
            p_vals[ethnicity2] = test_stats[1]

        except:
            statistics[ethnicity2] = np.nan
            p_vals[ethnicity2] = np.nan
    p_dict[ethnicity] = p_vals
    stats_dict[ethnicity] = statistics
```

In [31]:

```
p_to_plot = np.transpose(np.array([list(v.values()) for v in p_dict.values()]
stat_to_plot = np.transpose(np.array([list(v.values()) for v in stats_dict.va
X_vars =  np.array([list(v.keys()) for v in p_dict.values()])[0]
y_vars = np.array(list(p_dict.keys()))
```

In [32]:

```
fig, ax = plt.subplots(figsize=(15,15), dpi=300)
im = ax.imshow(p_to_plot, cmap='OrRd') # cmap='binary_r'
# We want to show all ticks...
ax.set_xticks(np.arange(len(X_vars)))
ax.set_yticks(np.arange(len(y_vars)))
# ... and label them with the respective list entries
ax.set_xticklabels(X_vars)
ax.set_yticklabels(y_vars)

# Rotate the tick labels and set their alignment.
plt.setp(ax.get_xticklabels(), rotation=45, ha="right",
         rotation_mode="anchor")

# Loop over data dimensions and create text annotations.
for i in range(len(y_vars)):
    for j in range(len(X_vars)):
        p_value_text = round(p_to_plot[i, j],2)

        if math.isnan(p_value_text):
            p_value_text = '= N/A'
            text_color_red = False
        elif p_value_text == 1.0:
            p_value_text = '> 0.99'
            text_color_red = False
        elif p_value_text == 0.0:
            p_value_text = '< 0.01'
            text_color_red = True
        else:
            if p_value_text < 0.05:
                text_color_red = True
            else:
                text_color_red = False
            p_value_text = '= {}'.format(p_value_text)

        stat_value_text = round(stat_to_plot[i, j],2)
        if math.isnan(stat_value_text):
            stat_value_text = 'N/A'
        text_to_plot = 'Statistic = {stat}\n p {p}'.format(stat=stat_value_te

        if p_value_text != 'N/A' and text_color_red:
            text = ax.text(j, i, text_to_plot,
                           ha="center", va="center", color="r")
        else:
            text = ax.text(j, i, text_to_plot,
                           ha="center", va="center", color="black")


ax.set_xlabel("Reference Population", size=12)
ax.set_ylabel("Test Population", size=12)
ax.set_title("Loss of Function Allele Frequency Mann-Whitney Results", size=1
fig.tight_layout()
plt.show()
fig.savefig('LoF_heatmap.svg', format='svg')
```

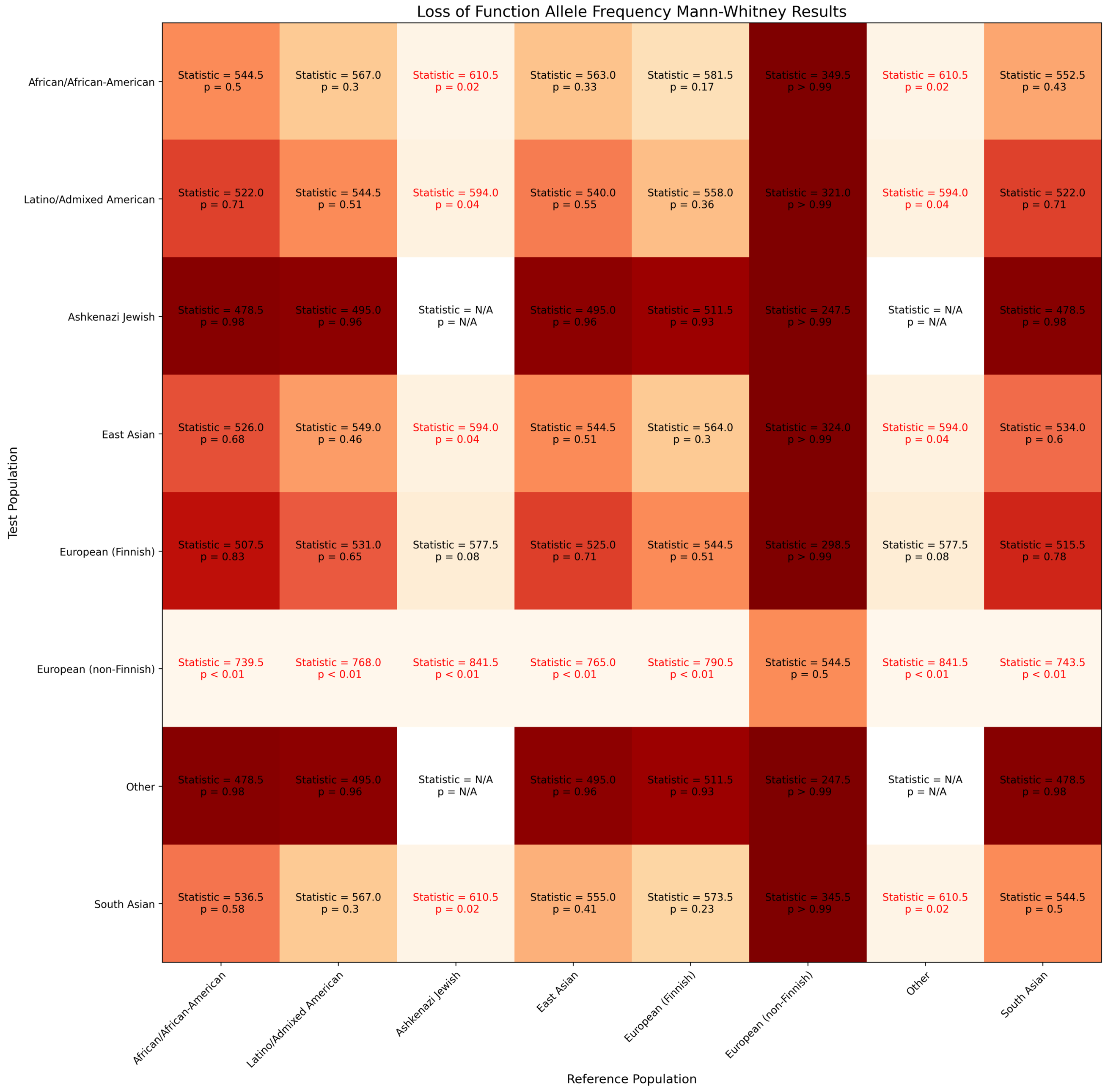
Loss of Function Allele Frequency Mann-Whitney Results
Test Population
Reference Population
African/African-American
Latino/Admixed American
Ashkenazi Jewish
East Asian
European (Finnish)
European (non-Finnish)
Other
South Asian
Statistic = 544.5 p = 0.5
Statistic = 567.0 p = 0.3
Statistic = 610.5 p = 0.02
Statistic = 563.0 p = 0.33
Statistic = 581.5 p = 0.17
Statistic = 349.5 p > 0.99
Statistic = 610.5 p = 0.02
Statistic = 552.5 p = 0.43
Statistic = 522.0 p = 0.71
Statistic = 544.5 p = 0.51
Statistic = 594.0 p = 0.04
Statistic = 540.0 p = 0.55
Statistic = 558.0 p = 0.36
Statistic = 321.0 p > 0.99
Statistic = 594.0 p = 0.04
Statistic = 522.0 p = 0.71
Statistic = 478.5 p = 0.98
Statistic = 495.0 p = 0.96
Statistic = N/A p = N/A
Statistic = 495.0 p = 0.96
Statistic = 511.5 p = 0.93
Statistic = 247.5 p > 0.99
Statistic = N/A p = N/A
Statistic = 478.5 p = 0.98
Statistic = 526.0 p = 0.68
Statistic = 549.0 p = 0.46
Statistic = 594.0 p = 0.04
Statistic = 544.5 p = 0.51
Statistic = 564.0 p = 0.3
Statistic = 324.0 p > 0.99
Statistic = 594.0 p = 0.04
Statistic = 534.0 p = 0.6
Statistic = 507.5 p = 0.83
Statistic = 531.0 p = 0.65
Statistic = 577.5 p = 0.08
Statistic = 525.0 p = 0.71
Statistic = 544.5 p = 0.51
Statistic = 298.5 p > 0.99
Statistic = 577.5 p = 0.08
Statistic = 515.5 p = 0.78
Statistic = 739.5 p < 0.01
Statistic = 768.0 p < 0.01
Statistic = 841.5 p < 0.01
Statistic = 765.0 p < 0.01
Statistic = 790.5 p < 0.01
Statistic = 544.5 p = 0.5
Statistic = 841.5 p < 0.01
Statistic = 743.5 p < 0.01
Statistic = 478.5 p = 0.98
Statistic = 495.0 p = 0.96
Statistic = N/A p = N/A
Statistic = 495.0 p = 0.96
Statistic = 511.5 p = 0.93
Statistic = 247.5 p > 0.99
Statistic = N/A p = N/A
Statistic = 478.5 p = 0.98
Statistic = 536.5 p = 0.58
Statistic = 567.0 p = 0.3
Statistic = 610.5 p = 0.02
Statistic = 555.0 p = 0.41
Statistic = 573.5 p = 0.23
Statistic = 345.5 p > 0.99
Statistic = 610.5 p = 0.02
Statistic = 544.5 p = 0.5
